# Evaluation of the diagnostic value of YiDiXie™-SS, YiDiXie™-HS and YiDiXie™-D in esophageal cancer

**DOI:** 10.1101/2024.09.15.24313696

**Authors:** Xutai Li, Zhenjian Ge, Peng Liao, Chen Sun, Wenkang Chen, Yingqi Li, Shengjie Lin, Pengwu Zhang, Wuping Wang, Siwei Chen, Yutong Wu, Huimei Zhou, Wei Li, Jing Du, Fangting Zhang, Yongqing Lai

## Abstract

**Background:** Esophageal cancer is a serious threat to human health and causes a heavy economic burden. upper gastrointestinal imaging(UGI imaging), enhanced CT, Fecal occult blood Tumor markers such as test(FOBT) and CEA, CA125 and CA19-9 are widely used in the screening or preliminary diagnosis of esophageal cancer. However, false-positive results of UGI imaging, enhanced CT, FOBT, CEA, CA125, and CA19-9 can lead to misdiagnosis and erroneous esophagoscopy, while their false-negative results can lead to missed diagnosis and delayed treatment. It is urgent to find a convenient, economical and non-invasive diagnostic method to reduce the false positive rate and false negative rate of UGI imaging. The objective of this study was to evaluate the diagnostic value of YiDiXie™-SS, YiDiXie™ -HS and YiDiXie™-D in esophageal cancer.

**Patients and methods:** This study included 164 subjects (malignant group, n=105; Benign group, n=59 cases). The remaining serum samples of the subjects were collected and the sensitivity and specificity of the YiDiXie™-SS, YiDiXie™ -HS and YiDiXie™-D were evaluated using the YiDiXie™all-cancer detection kit.

**Results:** The sensitivity of YiDiXie™-SS was 99.0% (95% CI: 94.8% - 100%) and its specificity was 61.0% (95% CI: 48.3% - 72.4%). This means that YiDiXie™-SS has an extremely high sensitivity and relatively high specificity in esophageal tumors.YiDiXie ™-HS has a sensitivity of 92.3% (95% CI: 85.7% - 96.1%) and a specificity of 86.4% (95% CI: 75.5% - 93.0%). This means that YiDiXie™-HS has high sensitivity and specificity in esophageal tumors.YiDiXie™-D has a sensitivity of 83.8% (95% CI: 75.6% - 89.6%) and a specificity of 93.2% (95% CI: 83.8% - 97.3%). This means that YiDiXie™-D has relatively high sensitivity and very high specificity in esophageal tumors. YiDiXie™-SS significantly reduced the false-positive rate of UGI imaging, CT, FOBT, CEA, CA125, CA19-9 with essentially no increase in malignancy leakage.YiDiXie™-HS significantly reduced the false-negative rate of UGI imaging, CT, FOBT, CEA, CA125, CA19-9. YiDiXie™-D significantly reduces the false positive rate of UGI imaging, CT, FOBT, CEA, CA125, CA19-9. YiDiXie™-D significantly reduces the false negative rate of UGI imaging, CT, FOBT, CEA, CA125, CA19-9, while maintaining a high level of specificity.

**Conclusion:** YiDiXie™-SS has very high sensitivity and relatively high specificity in esophageal tumors.YiDiXie™-HS has high sensitivity and high specificity in esophageal tumors.YiDiXie ™ -D has relatively high sensitivity and very high specificity in esophageal tumors. YiDiXie™-SS significantly reduced the false-positive rates of UGI imaging, CT, FOBT, CEA, CA125, and CA19-9 with essentially no increase in delayed treatment for esophageal cancer. YiDiXie ™ -HS substantially reduced the false-negative rates of UGI imaging, CT, FOBT, CEA, CA125, and CA19-9. YiDiXie ™ -D significantly reduces the false-positive rate of UGI imaging, CT, FOBT, CEA, CA125, CA19-9, or significantly reduces their false-negative rates while maintaining high specificity. YiDiXie™ tests have an important diagnostic value in esophageal cancer, and are expected to solve the problems of “high false-positive rate” and “high false-negative rate” of UGI imaging, CT, FOBT, CEA, CA125, CA19-9.

**Clinical trial number:** ChiCTR2200066840.

## INTRODUCTION

Esophageal carcinoma is among the most prevalent malignant tumors. According to the most recent data, 510,000 new instances of esophageal cancer would be diagnosed globally in 2022, with 440,000 additional fatalities^1^. Esophageal cancer is the seventh major cause of cancer-related death and the eleventh most frequent cancer globally. Patients with esophageal cancer had a lower survival rate when their tumor was aggressive, had a lymph node, or had distant metastases. The total 5-year survival rate is 15-25%, with early diagnosis yielding better results than late diagnosis^2^. Poor prognosis in individuals with esophageal cancer is frequently related with an advanced (metastatic) diagnosis and the cancer’s ability to metastasis, even if the tumor is merely superficial^3^. The prognosis for esophageal cancer is favorable only in the very early stages^4^, while esophagectomy remains the primary treatment for locally advanced cancer^5^. Therefore, esophageal cancer is a serious threat to human healt.

Upper gastrointestinal imaging (UGI) and enhanced CT are commonly used to screen for and diagnose esophageal cancer. On the one hand, UGI imaging and enhanced CT can generate a high percentage of false positives. The barium meal research had a positive predictive value of 42%^6^. Barium meal X-rays correctly diagnose esophageal cancer at 83.8% accuracy^7^. The false positive rate of enhanced CT in evaluating esophageal cancer is 5-10%^8-10^, with a positive predictive value of only 31.3%^10^. When UGI imaging and enhanced CT results are good, patients are frequently scheduled for esophagoscopy. False positive results from UGI imaging and enhanced CT mean that the patient has received an unnecessarily expensive and invasive esophagoscopy, and the patient will have to bear the adverse consequences of mental distress, expensive examination costs, and examination injuries. Therefore, it is urgent to find a convenient, economical and non-invasive diagnostic method to reduce the false positive rate of UGI imaging and enhanced CT.

On the other hand, Enhanced CT and UGI imaging can result in a significant percentage of false negatives. The false negative rate for esophageal cancer using X-ray barium meal was 15 – 55%^7,11-12^. The enhanced CT assessment of esophageal cancer has a 40% false-negative rate^8^. Up to 78% of enhanced CT scans for esophageal cancer were falsely negative^10^. Patients often have observation and routine follow-up when results from enhanced CT and UGI imaging are negative. False negative results from UGI imaging and enhanced CT mean that the malignant tumor is misdiagnosed as benign disease, which can lead to delayed treatment, progression of the malignant tumor, and possibly advanced stage development. Therefore, patients will have to bear the adverse consequences of poor prognosis, high treatment costs, poor quality of life, and short survival. Therefore, it is urgent to find a convenient, economical and non-invasive diagnostic method to reduce the false negative rate of UGI imaging and enhanced CT.

Furthermore, tumor markers like CEA, CA125 and CA19-9, as well as the fecal occult blood test (FOBT), are frequently employed in the screening process for esophageal cancer. On the one hand, a lot of false positive results can be obtained from procedures like UGI imaging, enhanced CT, FOBT, CEA, CA125 CA19-9, etc. 32.5% of CEA+CA199 combination detection cases resulted in false positives^13^. Patients often get an esophagoscopy if UGI imaging is positive. False positive results, like UGI imaging, indicate that the patient underwent an intrusive and costly esophagoscopy without necessity. As a result, the patient will suffer from psychological distress, high examination fees, and examination-related injuries. Therefore, it is urgent to find a convenient, economical and non-invasive diagnostic method to reduce the false positive rate of UGI imaging.

On the other side, a lot of false negative results can be obtained from procedures like FOBT, CEA, CA125 CA19-9, etc. 80% of FOBT false negative results are associated with esophageal cancer^14-15^. The percentages of false negative results for CEA and CA19-9 in cases of esophageal cancer were 60-75% and 66-82%, respectively^16-17^. The negative predictive values were 61.72% and 54.94%, respectively^16^. Up to 90% of CA125 tests result in erroneous negative results^18^. In the event that UGI imaging is negative, patients are typically monitored and routinely checked on. False negative results such as UGI imaging mean that the malignant tumor is misdiagnosed as a benign disease, which may lead to delayed treatment, the progression of the malignant tumor, and possibly even the advanced stage. Therefore, patients will have to bear the adverse consequences of poor prognosis, high treatment costs, poor quality of life, and short survival. Therefore, it is urgent to find a convenient, economical and non-invasive diagnostic method to reduce the false negative rate of UGI imaging.

Based on the detection of novel tumor markers of miRNA in serum, Shenzhen Kerida Health Technology Co., Ltd. has developed an in vitro diagnostic test product: YiDiXie ™ all-cancer test (hereinafter referred to as “the YiDiXie™ test”). The YiDiXie ™ test, which can detect multiple cancer types with just 200 microliters of whole blood or 100 microliters of serum at a time^19^. The “YiDiXie ™ test” includes three products with different performance: YiDiXie™-HS, YiDiXie™-SS, and YiDiXie™-D^19^.

The aim of this study is to evaluate the diagnostic value of YiDiXie™-SS, YiDiXie™-HS and YiDiXie™-D in esophageal cancer.

## PATIENTS AND METHODS

### Study design

This study is part of the sub-study “Assessment of the YiDiXie ™ test as an adjunct diagnostic tool across various tumors” within the SZ-PILOT project (ChiCTR2200066840).

SZ-PILOT is a forward-looking, observational, single-center trial (ChiCTR2200066840). For this investigation, 0.5 ml of leftover serum from participants who had consented to donate residual samples during their physical exams or admissions were utilized.

The study was conducted with a blinded design. Laboratory technicians performed the YiDiXie ™ test without access to the participants’ clinical information. Results were then reviewed by KeRuiDa laboratory staff. Similarly, the clinical professionals assessing the participants’ clinical data were unaware of the YiDiXie™ test results.

The study received approval from the Ethics Committee of Shenzhen Hospital at Peking University and was conducted in adherence to the International Conference on Harmonization (ICH) guidelines for the Quality Management of Pharmaceutical Clinical Trials and the Declaration of Helsinki.

### Participants

The study included patients with esophageal cancer and colorectal benign disease who had UGI imaging, enhanced CT, FOBT, CEA, CA125 and CA19-9 test results. Both groups were enrolled individually, and all participants who met the inclusion criteria were included indefinitely.

Patients with “suspected (solid or blood) malignancy” who completed the paninformed consent form to provide remaining samples were initially included in this study. Subjects having a postoperative pathological diagnosis of “malignancy” were placed in the malignant group, whereas those with a postoperative pathological diagnosis of “benign disease” were put in the benign group. This investigation omitted pathology outcomes that were benign or malignant. The benign group also included healthy exam patients with esophagoscopy results. Previous studies by our study group^19^ employed samples from both the malignant tumor group and the benign group of healthy medical examiners.

Participants who failed serum sample quality tests prior to the YiDiXie ™ test were eliminated from the study. Refer to our study group’s earlier studies for the precise enrollment and exclusion scenario^19^.

### Sample collection, processing

There was no need for additional blood sampling because the serum samples used in this inquiry were from serum left over from a normal consultation. For the YiDiXie ™ test, the Medical Laboratory removed approximately 0.5 cc of serum from each participant and stored it at -80°C.

### The YiDiXie test

Shenzhen KeRuiDa Health Technology Co. developed and manufactures the YiDiXie ™ all-cancer detection kit, an in vitro diagnostic kit for fluorescent quantitative PCR machines used to perform the YiDiXie™ test^19^. It detects cancer in the subject’s body by measuring the expression levels of several miRNA biomarkers in the serum^19^. It preserves specificity and boosts sensitivity for a wide range of malignancies by combining these independent assays in a real-time testing format and predefining appropriate parameters for each miRNA biomarker to ensure that each miRNA marker is highly specific^19^.

The YiDiXie™ test includes three unique tests: YiDiXie ™ -HS, YiDiXie ™ -SS, and YiDiXie ™ -D. YiDiXie ™ -HS was developed with a focus on specificity and sensitivity^19^. YiDiXie™-SS significantly increased the number of miRNA assays, resulting in extremely high sensitivity for all clinical stages of malignant tumors^19^. YiDiXie™-D considerably raises the diagnostic threshold for individual miRNA testing, resulting in high specificity for all types of cancer^19^.

To perform the YiDiXie ™ test, follow the instructions provided. Refer to our prior work^19^ for comprehensive procedures.

Laboratory technicians at Shenzhen KeRuiDa Health Technology Co., Ltd. analyze raw test findings and determine if the YiDiXie ™ test is “positive” or “negative”^19^.

### Extraction of clinical data

The clinical, pathological, laboratory, and imaging data for this inquiry were sourced from the patients’ inpatient medical records or physical examination reports. Clinical staging was performed by trained physicians using the AJCC staging manual (7th or 8th edition)^20-21^.

### Statistical analyses

Descriptive statistics were used to record baseline and demographic data. For continuous variables, the mean, standard deviation (SD) or standard error (SE), median, first quartile (Q1), third quartile (Q3), minimum and maximum values were calculated. For categorical variables, the number and percentage of subjects in each category were calculated. For a number of metrics, the 95% confidence intervals (CIs) were calculated using the Wilson (score) technique.

## RESULTS

### Participant disposition

There were 164 individuals total (105 malignant and 59 benign). The 164 research participants’ clinical and demographic details are listed in Table 1.

**Table 1.** Participants’ demographic and clinical manifestation.

|  | Malignant (n=105) | Benign (n=59) | Total (N=164) |
| --- | --- | --- | --- |
| Age, years |  |  |  |
| Mean (SD) | 64.9 (8.20) | 55.9 (9.82) | 61.6 (9.79) |
| Median (Q1,Q3) | 65 (59, 69) | 56 (51, 65) | 62 (56, 68) |
| Min, max | 40, 90 | 30, 71 | 30, 90 |
| Age, group, n (%) |  |  |  |
| < 50 | 4 (3.8) | 11 (18.6) | 15 (9.1) |
| ≥ 50 | 101 (96.2) | 48 (81.4) | 149 (90.9) |
| < 65 | 50 (47.6) | 44 (74.6) | 94 (57.3) |
| ≥ 65 | 55 (52.4) | 15 (25.4) | 70 (42.7) |
| Sex, n (%) |  |  |  |
| Female | 20 (19.0) | 19 (32.2) | 39 (23.8) |
| Male | 85 (81.0) | 40 (67.8) | 125 (76.2) |
| Body mass index (kg/m<sup>2</sup>) |  |  |  |
| n | 98 | 50 | 148 |
| Mean (SD) | 21.9 (3.71) | 24.3 (3.41) | 22.7 (3.77) |
| Median (Q1,Q3) | 21.5 (19.2, 24.2) | 23.9 (21.5, 26.4) | 22.4 (19.8, 24.8) |
| Min, max | 13.0 33.3 | 17.3 34.2 | 13.0 34.2 |
| Body mass index category, n (%) |  |  |  |
| Underweight | 19 (18.1) | 1 (1.7) | 20 (12.2) |
| Benign | 51 (48.6) | 24 (40.7) | 75 (45.7) |
| Overweight | 22 (21.0) | 19 (32.2) | 41 (25.0) |
| Obese | 6 (5.7) | 6 (10.2) | 12 (7.3) |
| Missing | 7 (6.7) | 9 (15.3) | 16 (9.8) |
| AJCC clinical stage |  |  |  |
| Stage 0 | 2 (1.9) |  | 2 (1.9) |
| Stage I | 10 (9.5) |  | 10 (9.5) |
| Stage II | 33 (31.4) |  | 33 (31.4) |
| Stage III | 43 (41.0) |  | 43 (41.0) |
| Stage IV | 8 (7.6) |  | 8 (7.6) |
| Missing | 9 (8.6) |  | 9 (8.6) |

Q1,Q3, first quartile, third quartile; SD, standard deviation.

The clinical and demographic features of the two groups were similar (Table 1). Aged 61.6 (9.79) years on average (standard deviation), 23.8% (39/164) of the population was female.

### Diagnostic performance of YiDiXie™-SS

As shown in Table 2, the sensitivity of YiDiXie™ -SS was 99.0% (95% CI: 94.8% - 100%) and its specificity was 61.0% (95% CI: 48.3% - 72.4%). This means that YiDiXie ™ -SS has very high sensitivity and relatively high specificity in esophageal tumors.

**Table 2.** Performance of YiDiXie™-SS.

|  | Malignant | Benign | Total |
| --- | --- | --- | --- |
|  | 105 | 59 | 164 |
| Positive | 104 | 23 | 127 |
| Negative | 1 | 36 | 37 |

Two-sided 95% Wilson confidence intervals were calculated. SEN, Sensitivity. SPE, Specificity.

### Diagnostic performance of YiDiXie™-HS

As shown in Table 3, the sensitivity of YiDiXie™ -HS was 92.3% (95% CI: 85.7% - 96.1%) and its specificity was 86.4% (95% CI: 75.5% - 93.0%). This means that YiDiXie ™ -HS has high sensitivity and high specificity in esophageal tumors.

**Table 3.** Performance of YiDiXie™-HS.

|  | Malignant | Benign | Total |
| --- | --- | --- | --- |
|  | 105 | 59 | 164 |
| Positive | 97 | 8 | 105 |
| Negative | 8 | 51 | 59 |

Two-sided 95% Wilson confidence intervals were calculated. SEN, Sensitivity. SPE, Specificity.

### Diagnostic performance of YiDiXie™-D

As shown in Table 4, the sensitivity of YiDiXie™ -D was 83.8% (95% CI: 75.6% - 89.6%) and its specificity was 93.2% (95% CI: 83.8% - 97.3%). This means that YiDiXie ™ -D has relatively high sensitivity and very high specificity in esophageal tumors.

**Table 4.** Performance of YiDiXie™-D.

|  | Malignant | Benign | Total |
| --- | --- | --- | --- |
|  | 105 | 59 | 164 |
| Positive | 88 | 4 | 92 |
| Negative | 17 | 55 | 72 |

Two-sided 95% Wilson confidence intervals were calculated. SEN, Sensitivity. SPE, Specificity.

### Diagnostic performance of YiDiXie ™ -SS in UGI imaging, CT, FOBT, CEA, CA125, CA19-9 positive patients

In order to solve the challenge of high false-positive rate of UGI imaging, CT, FOBT, CEA, CA125, CA19-9, YiDiXie™-SS was applied to UGI imaging, CT, FOBT, CEA, CA125, CA19-9 positive patients.

As shown in Table 5, YiDiXie ™-SS significantly reduced the false-positive rates of UGI imaging, CT, FOBT, CEA, CA125, and CA19-9 with essentially no increase in malignancy leakage.

**Table 5.** Performance of different Items.

| Items | Total N | Positive | Sensitivity % (95% CI)<sup>a</sup> | Total N | Negative | Specificity % (95% CI)<sup>b</sup> |
| --- | --- | --- | --- | --- | --- | --- |
| YiDiXie™-SS in UGII-positive | 29 | 28 | 96.6% (82.8% - 99.8%) | 5 | 3 | 60.0% (23.1% - 92.9%) |
| YiDiXie™-HS in UGII-negative | 9 | 8 | 88.9% (56.5% - 99.4%) | 5 | 3 | 60.0% (23.1% - 92.9%) |
| YiDiXie™-D in UGII-positive | 29 | 24 | 82.8% (65.5% - 92.4%) | 5 | 5 | 100% (56.6% - 100%) |
| YiDiXie™-D in UGII-negative | 9 | 7 | 77.8% (45.3% - 96.1%) | 5 | 4 | 80.0% (37.6% - 99.0%) |
| YiDiXie™-SS in CT-positive | 78 | 77 | 98.7% (93.1% - 99.9%) | 4 | 3 | 75.0% (30.1% - 98.7%) |
| YiDiXie™-HS in CT-negative | 22 | 20 | 90.9% (72.2% - 98.4%) | 6 | 3 | 50.0% (18.8% - 81.2%) |
| YiDiXie™-D in CT-positive | 78 | 66 | 84.6% (75.0% - 91.0%) | 4 | 4 | 100% (51.0% - 100%) |
| YiDiXie™-D in CT-negative | 22 | 18 | 81.8% (61.5% - 92.7%) | 6 | 5 | 83.3% (43.6% - 99.1%) |
| YiDiXie™-SS in FOBT-positive | 2 | 2 | 100% (17.8% - 100%) | 1 | 0 | 0 (5.1% - 100%) |
| YiDiXie™-HS in FOBT-negative | 32 | 29 | 90.6% (75.8% - 96.8%) | 5 | 4 | 80.0% (37.6% - 99.0%) |
| YiDiXie™-D in FOBT-positive | 2 | 2 | 100% (17.8% - 100%) | 1 | 1 | 100% (5.1% - 100%) |
| YiDiXie™-D in FOBT-negative | 32 | 25 | 78.1% (61.2% - 89.0%) | 5 | 5 | 100% (56.6% - 100%) |
| YiDiXie™-SS in CEA-positive | 17 | 17 | 100% (81.6% - 100%) | 3 | 2 | 66.7% (11.8% - 98.3%) |
| YiDiXie™-HS in CEA-negative | 81 | 76 | 93.8% (86.4% - 97.3%) | 55 | 48 | 87.3% (76.0% - 93.7%) |
| YiDiXie™-D in CEA-positive | 17 | 14 | 82.4% (59.0% - 93.8%) | 3 | 3 | 100% (43.9% - 100%) |
| YiDiXie™-D in CEA-negative | 81 | 72 | 88.9% (80.2% - 94.0%) | 55 | 52 | 94.5% (85.1% - 98.5%) |
| YiDiXie™-SS in CA125-positive | 6 | 6 | 100% (61.0% - 100%) | 1 | 1 | 100% (5.1% - 100%) |
| YiDiXie™-HS in CA125-negative | 88 | 82 | 93.2% (85.9% - 96.8%) | 29 | 23 | 79.3% (61.6% - 90.2%) |
| YiDiXie™-D in CA125-positive | 6 | 5 | 83.3% (43.6% - 99.1%) | 1 | 1 | 100% (5.1% - 100%) |
| YiDiXie™-D in CA125-negative | 88 | 78 | 88.6% (80.3% - 93.7%) | 29 | 28 | 96.6% (82.8% - 99.8%) |
| YiDiXie™-SS in CA19-9-positive | 5 | 5 | 100% (56.6% - 100%) | 2 | 0 | 0 (0 - 82.2%) |
| YiDiXie™-HS in CA19-9-negative | 92 | 85 | 92.4% (85.1% - 96.3%) | 54 | 47 | 87.0% (75.6% - 93.6%) |
| YiDiXie™-D in CA19-9-positive | 5 | 5 | 100% (56.6% - 100%) | 2 | 2 | 100% (17.8% - 100%) |
| YiDiXie™-D in CA19-9-negative | 92 | 78 | 84.8% (76.1% - 90.7%) | 54 | 51 | 94.4% (84.9% - 98.5%) |

UGII, UGI imaging. CI, confidence interval. <sup>ab</sup> Two-sided 95% Wilson confidence intervals were calculated.

### Diagnostic performance of YiDiXie ™ -HS in UGI imaging, CT, FOBT, CEA, CA125, CA19-9 negative patients

In order to solve the difficult problem of high false-negative rate of UGI imaging, CT, FOBT, CEA, CA125, and CA19-9, YiDiXie™-HS was applied to patients with negative UGI imaging, CT, FOBT, CEA, CA125, and CA19-9.

As shown in Table 5, YiDiXie™-HS substantially reduced the false-negative rates of UGI imaging, CT, FOBT, CEA, CA125, and CA19-9.

### Diagnostic performance of YiDiXie™-D in UGI imaging, CT, FOBT, CEA, CA125, CA19-9 positive patients

In order to further reduce the false-positive rate of UGI imaging, CT, FOBT, CEA, CA125, and CA19-9, YiDiXie ™ -D, which has relatively high sensitivity and very high specificity, was therefore applied to UGI imaging, CT, FOBT, CEA, CA125, and CA19-9 positive patients.

As shown in Table 5, YiDiXie™-D substantially reduced the false-positive rates of UGI imaging, CT, FOBT, CEA, CA125, and CA19-9.

### Diagnostic performance of YiDiXie ™ -D in UGI imaging, CT, FOBT, CEA, CA125, CA19-9 negative patients

In order to reduce the false-negative rate of UGI imaging, CT, FOBT, CEA, CA125, and CA19-9 while maintaining a high degree of specificity, YiDiXie™-D, which has a relatively high sensitivity and a very high degree of specificity, was applied to patients negative for UGI imaging, CT, FOBT, CEA, CA125, and CA19-9.

As shown in Table 5, YiDiXie™-D significantly reduced the false-negative rate of UGI imaging, CT, FOBT, CEA, CA125, and CA19-9 while maintaining a high specificity.

## DISCUSSION

### Clinical significance of YiDiXie™-SS in patients with positive UGI imaging and other indicators

For positive patients, such as those with UGI imaging, the sensitivity and specificity of subsequent diagnostic procedures are critical. evaluating the contradiction between sensitivity and specificity entails evaluating “the harm of missing diagnosis of malignant tumors” against “the harm of misdiagnosis of benign tumors.” In general, when UGI imaging is positive, esophagoscopy is used instead of aggressive surgery. False positives, such as UGI imaging, do not result in catastrophic repercussions such as substantial surgical trauma, organ removal, or loss of function. In this method, for positive patients, such as UGI imaging, the “harm of missed diagnosis of malignant tumor” is significantly more than the “harm of misdiagnosis of benign tumor.” Therefore, YiDiXie ™ -SS with extremely high sensitivity but slightly low specificity was selected to reduce false positive rates such as UGI imaging.

As shown in Table 5, YiDiXie™-SS significantly reduced the false-positive rates of UGI imaging, CT, FOBT, CEA, CA125, and CA19-9 with essentially no increase in malignancy leakage. YiDiXie ™ -SS minimizes erroneous colonoscopy for benign colorectal disease while without increasing the likelihood of missed malignant cancers.

In other findings, YiDiXie ™ -SS considerably reduced mental discomfort, expensive examination costs, and examination injuries in false positive patients with UGI imaging, enhanced CT, FOBT, CEA, CA125 and CA19-9 without significantly increasing the delayed treatment of malignant tumors. YiDiXie™-SS is clinically significant and has a wide range of applications.

### Clinical significance of YiDiXie™-HS in patients with negative UGI imaging and other indicators

The sensitivity and specificity of additional diagnostic techniques are critical for patients with negative UGI imaging and other signs. Weighing the conflict between the “harm of malignant tumor underdiagnosis” and the “harm of benign disease misdiagnosis” is the equivalent of weighing the conflict between sensitivity and specificity. Increased false-negative rates translate into an increased number of malignant tumors being underdiagnosed, which delays treatment, causes the tumor to develop, and can even reach advanced stages. Patients will consequently have to deal with the negative effects of a bad prognosis, a brief surviving time, a low quality of life, and expensive therapy. A higher false-positive rate translates into more benign diseases being misdiagnosed, which will needlessly result in an invasive and costly colonoscopy. Patients must therefore deal with the effects of psychological distress, costly testing, and injury. Therefore, YiDiXie™-HS, with its high sensitivity and specificity, was chosen to reduce the false negative rate of UGI imaging and other indicators.

As shown in Table 5, YiDiXie™-HS substantially reduced the false-negative rates of UGI imaging, CT, FOBT, CEA, CA125, and CA19-9. YiDiXie™-HS minimizes false negative missed cancers, including UGI imaging.

YiDiXie ™ -HS dramatically improves bad outcomes for patients with false negative misdiagnoses, including poor prognosis, high treatment costs, poor quality of life, and short survival due to UGI imaging. YiDiXie ™ -HS is clinically significant and has a wide range of application opportunities.

### Clinical significance of YiDiXie™-D in patients with esophageal tumors

For patients with esophageal tumors, YiDiXie™-D, which has relatively high sensitivity and very high specificity, can be used to further reduce the false-positive rate or significantly reduce the false-negative rate of UGI imaging, CT, FOBT, CEA, CA125, and CA19-9 while maintaining high specificity.

As shown in Table 5, YiDiXie™-D substantially reduced the false-positive rates of UGI imaging, CT, FOBT, CEA, CA125, and CA19-9. YiDiXie ™ -D significantly reduced the false-negative rates of UGI imaging, CT, FOBT, CEA, CA125, and CA19-9 while maintaining a high degree of specificity.

The above results imply that YiDiXie ™ -D further reduces the risk of incorrectly performing esophagoscopy. Therefore, YiDiXie™-D meets the clinical needs well and has important clinical significance and wide application prospects.

### YiDiXie™ tests are expected to solve two challenges of esophageal cancer

First of all, YiDiXie™-SS reduces work pressure for digestive endoscopy physicians and promotes rapid diagnosis and treatment of malignant tumors that were previously postponed. When UGI imaging is positive, the patient is often evaluated via esophagoscopy. The amount of digestive endoscopists determines whether or not these esophagoscopies can be finished on schedule. In many regions of the world, bookings might take months or even years. This unavoidably delays the treatment of malignant tumors among them, therefore it is not uncommon for positive patients with UGI imaging who are awaiting colonoscopy to experience malignant tumor progression or even distant metastases. As shown in Table 5, YiDiXie™ -SS significantly reduced the false-positive rates of UGI imaging, CT, FOBT, CEA, CA125, and CA19-9 with essentially no increase in malignancy leakage. YiDiXie™-SS minimizes erroneous colonoscopy for benign colorectal disease while without increasing the likelihood of missed malignant cancers. Therefore, YiDiXie ™ -SS can greatly relieve the unnecessary work pressure of digestive endoscopists, and facilitate the timely diagnosis and treatment of esophageal cancer or other diseases that have been delayed.

Second, YiDiXie ™-HS dramatically lowers the chance of missing esophageal cancer. When UGI imaging yields negative results, esophageal cancer is usually temporarily ruled out. Because of the significant false negative rate of UGI imaging, many patients with esophageal cancer have postponed therapy. As shown in Table 5, YiDiXie ™ -HS substantially reduced the false-negative rates of UGI imaging, CT, FOBT, CEA, CA125, and CA19-9. As a result, YiDiXie ™-HS significantly reduces the probability of false-negative missed malignancies such as UGI imaging, facilitating timely diagnosis and treatment for patients with esophageal cancer who would otherwise have been delayed.

Again, YiDiXie ™ -D is expected to further address the challenges of “high false positive rate” and “high false negative rate”. As shown in Table 5, YiDiXie™-D substantially reduces the false-positive rates of FOBT, CEA, CA125, and CA19-9, or significantly reduces the false-negative rates of UGI imaging, CT, FOBT, CEA, CA125, and CA19-9 while maintaining a high degree of specificity. Thus, YiDiXie ™-D further reduces the risk of incorrectly performing esophagoscopy.

Final, the YiDiXie™ test enables “just-in-time diagnosis” for patients with esophageal cancer. On the one hand, the YiDiXie™ test requires only a tiny amount of blood, allowing patients to complete the diagnostic process without leaving their homes. The YiDiXie ™ test requires only 20 microliters of serum to complete, which is about the same amount as a drop of whole blood (a drop of whole blood is about 50 microliters and produces 20-25 microliters of serum)^19^. Taking into account the sample quality assessment test prior to testing and 2-3 repetitions, 0.2 ml of whole blood is sufficient to complete the YiDiXie™ test^19^. Ordinary subjects can use the finger blood collection needle to complete 0.2 ml finger blood collection at home, without the need for intravenous blood collection by medical personnel, and patients can complete the diagnosis process without leaving the house^19^.

The YiDiXie ™ test offers practically infinite diagnostic capabilities. Figure 1 depicts the fundamental flow diagram of the YiDiXie ™ test, which does not require a doctor, medical equipment, or blood collection. The YiDiXie™ test is independent of medical staff and institutions, allowing for practically unlimited testing capacity. The YiDiXie ™ test offers “just-in-time diagnosis” for patients with esophageal cancer, eliminating the anxiety of waiting for appointments.

**Figure 1.**
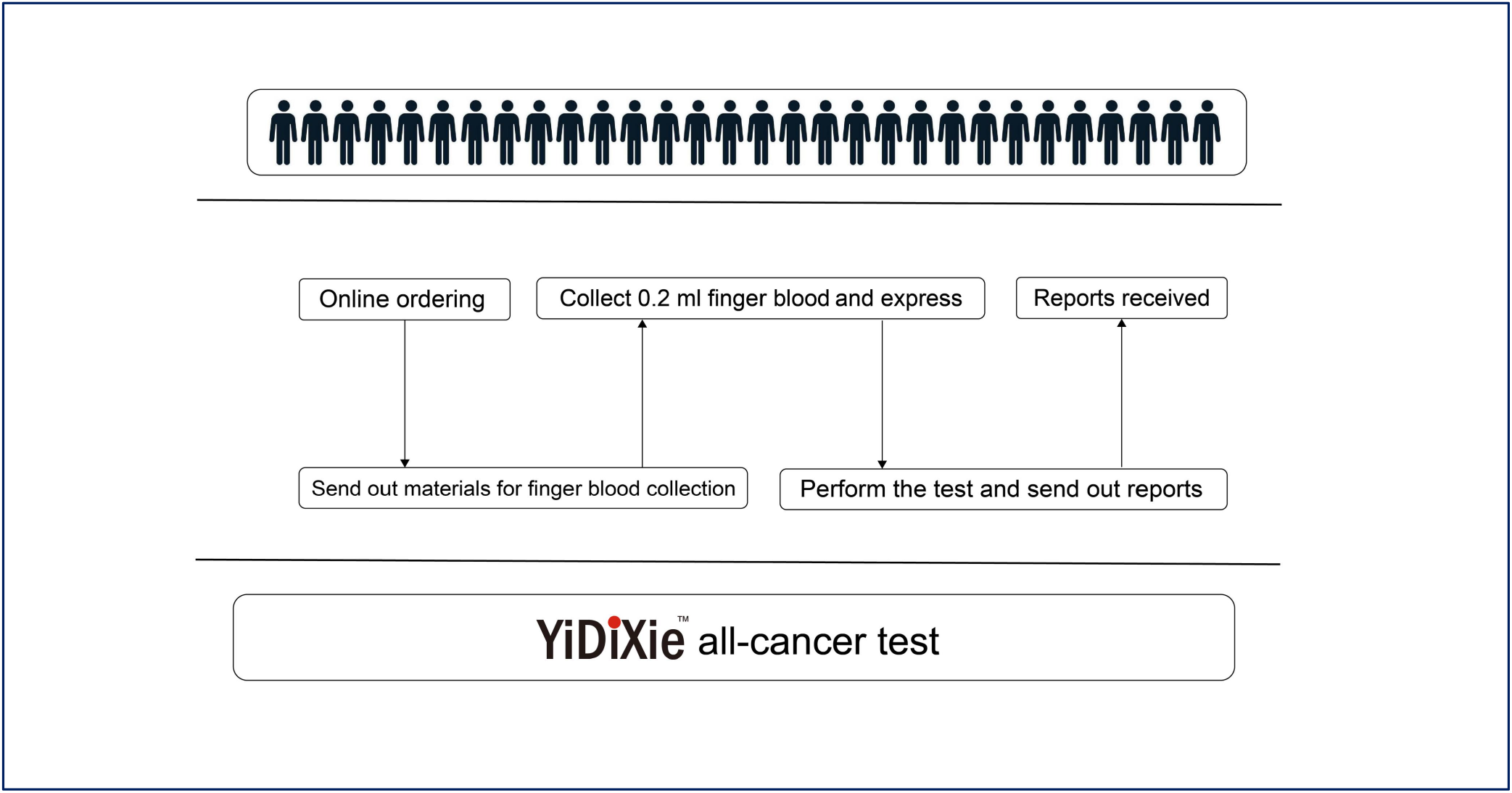
Basic flowchart of the YiDiXie™ test.

In short, the YiDiXie ™ test has important diagnostic value in esophageal cancer, and is expected to solve the two problems of “too high false positive rate such as UGI imaging” and “too high false negative rate such as UGI imaging” in esophageal cancer.

### Limitations of the study

First, the number of cases in this investigation was modest, necessitating future clinical trials with bigger sample sizes for further evaluation.

Second, this is a malignant tumor case-benign tumor control research in inpatients, therefore further cohort studies of the natural population of esophageal tumors are required for further evaluation.

Final, this was a single-center trial, which may have resulted in some bias. Additional multicenter studies are required for further evaluation.

## CONCLUSION

YiDiXie ™ -SS has very high sensitivity and relatively high specificity in esophageal tumors.YiDiXie ™-HS has high sensitivity and high specificity in esophageal tumors.YiDiXie ™ -D has relatively high sensitivity and very high specificity in esophageal tumors. YiDiXie ™ -SS significantly reduced the false-positive rates of UGI imaging, CT, FOBT, CEA, CA125, and CA19-9 with essentially no increase in delayed treatment for esophageal cancer. YiDiXie ™ -HS substantially reduced the false-negative rates of UGI imaging, CT, FOBT, CEA, CA125, and CA19-9. YiDiXie ™ -D significantly reduces the false-positive rate of UGI imaging, CT, FOBT, CEA, CA125, CA19-9, or significantly reduces their false-negative rates while maintaining high specificity. YiDiXie ™ tests have an important diagnostic value in esophageal cancer, and are expected to solve the problems of “high false-positive rate” and “high false-negative rate” of UGI imaging, CT, FOBT, CEA, CA125, CA19-9.

## Data Availability

All data produced in the present study are contained in the manuscript.

